# The causal role of adiposity in mental illness: A systematic review and meta-analysis of Mendelian randomization studies

**DOI:** 10.1101/2024.04.19.24304800

**Authors:** Min Gao, Paul J Doody, Dimitrios A Koutoukidis, Yebo Yu, Paul Aveyard, Richard Stevens, Padraig C Dixon

**Author notes:** **MG** and **PCD** are guarantors of the study.

## Abstract

**Importance:** Excess adiposity is associated with mental illnesses, but the causality of these associations remains equivocal.

**Objective:** To examine the causal effect of adiposity on mental illnesses by summarizing and assessing evidence from Mendelian randomization (MR) studies.

**Study Selection:** Searches were conducted on Embase, Medline, and Web of Science from database inception to January 8^th^ 2024.

**Data Extraction and Synthesis:** Studies using MR study designs that estimated adiposity measures including body mass index, abdominal adiposity, peripheral adiposity, or body composition in relation to mental illnesses were included. Study quality was assessed with a scoring system reflecting MR study guidelines. Data were pooled in meta-analyses using random-effects models. Subgroup analyses were conducted by sex.

**Main Outcomes and Measures:** Outcomes were the presence or severity of depression, anxiety, eating disorders, bipolar disorder, obsessive compulsive disorder, post-traumatic stress disorder, schizophrenia, and related psychotic disorders.

**Results:** Forty-one studies with 184 MR estimates were included in the systematic review; and 16 studies with 30 MR estimates were included in meta-analysis. Pooled estimates suggested that general adiposity was causally associated with depression (OR: 1.05, 95% CI, 1.00-1.10, p<0.001; I^2^=82.8%), though the effect size was modest and there was high heterogeneity. Subgroup difference by sex in the causal relationship was not **observed** (**p**=0.382). No evidence of causal associations was found between adiposity and other mental illnesses, though these analyses were characterized by high imprecision and heterogeneity.

**Conclusion and Relevance:** General adiposity appears to be causally associated with depression, suggesting psychological benefits of weight management. Evidence for causal associations between adiposity and other mental illnesses remains uncertain.

**Key Points:** *Question:* Is adiposity causally associated with mental illnesses?

*Findings:* In this systematic review of 41 Mendelian randomization studies, with 184 MR estimates, and meta-analysis of 30 MR estimates, general adiposity was causally associated with depression. There was no evidence of sex differences. Evidence of causal associations between adiposity and other mental illnesses was unclear.

*Meaning:* The findings suggest that higher adiposity can cause depression, which highlights the psychological importance of weight management.

## Introduction

One in eight people live with a mental illness worldwide, accounting for nearly one billion people ^1^. The association between adiposity and mental illness is well established. Approximately 20%-60% of people with obesity experience at least one form of mental illness ^2^. Adipose tissue is an endocrine organ that releases cytokines and other molecules that influence immune activation, insulin resistance, and lipid metabolism ^3^, which are theorized to cause, or exacerbate, mental illness ^4^. Psychological factors (e.g., weight stigma) may also explain the association ^5^. Evidence from cross-sectional and longitudinal studies indicates that body mass index (BMI) is strongly associated with depression ^6^, anxiety ^7^, eating disorders and serious mental illness (i.e., bipolar disorder, schizophrenia, and psychosis) ^8^. The distribution of adipose tissue in the body may also influence mental health. Abdominal adiposity produces a greater effect on systemic inflammation and homeostatic functions than other sites of excess adiposity and may have a stronger effect on health outcomes ^9^. Indeed, co-occurrence of mental illness with excess abdominal fat and metabolic disorders has been reported in epidemiological research ^10^.

However, previous evidence on the association between adiposity and mental illness may be confounded by reverse causation, bias, and residual confounding. Mental illness could lead to adiposity, partly through side effects of medication used to treat mental illness (i.e., weight gain), or through low motivation for weight control ^11^. Moreover, obesity co-occurs with many other risk factors which can confound the association between obesity and mental illness. For example, socioeconomic deprivation, social adversity, reduced physical activity, and unhealthy diet are associated with both obesity and mental illness ^12^. Therefore, assessing the causal association between adiposity and mental illness is challenging but is nonetheless crucial to understanding disease etiology and to develop effective diagnostic, therapeutic, and preventative interventions. Such knowledge also has major public health implications because, despite the introduction of psychological therapy and medication, many patients respond poorly to these current treatments^13,14^, and many psychotherapeutic agents increase the risk of weight gain ^15,16^.

Mendelian randomization (MR) analyses use genetic variants as instrumental variables, which - in principle - overcome the reverse causation and confounding that affects traditional or conventional observational study designs ^17^. Mendelian randomization permits a robust assessment of the causal relationship between adiposity and mental illness. Recent MR studies have explored the causal role of adiposity in multiple mental illnesses, but results are yet to be summarized and assessed. We conducted a systematic review and meta-analysis of MR studies to appraise and summarize evidence on causality between adiposity and mental illnesses.

## Methods

This systematic review was conducted in accordance with the Preferred Reporting Items for Systematic Reviews and Meta-Analysis (PRISMA) statement and the recent STROBE-MR guideline ^18,19^. The protocol for the systematic review was prospectively registered with the International Prospective Register of Systematic Reviews (PROSPERO) on 31^st^ May, 2022 (CRD42022336439).

### Search strategy

Searches were conducted on Embase, Medline, and Web of Science from database inception to January 8^th^ 2024, using search terms that included synonyms of “adiposity” AND “mental illness” AND “Mendelian randomization”. This study was designed to include causal evidence that performed analyses referred to as “Mendelian randomization” (or closely related methods such as “genetic instrumental variable regression”) as detailed in ***Supplementary material 1*** and ***Supplementary material 2*** . We included studies that investigated the causal associations of adiposity (e.g., body mass index, body fat percentage), abdominal adiposity (e.g., waist circumference, waist-to-hip ratio), peripheral adiposity (e.g., arm fat mass) and body composition (e.g., fat mass index) with depression, anxiety, eating disorders, bipolar disorder, obsessive compulsive disorder (OCD), post-traumatic stress disorder (PTSD), schizophrenia and other related disorders. The lead reviewer (MG) and one other reviewer (PJD, DAK, or YY) independently screened potentially eligible studies based on title and abstract, followed by full text where applicable.

### Data extraction

Data were extracted and entered in predefined data extraction tables independently by the lead reviewer (MG) and one of the other three reviewers (PJD, DAK, or YY) and finalized after discussion (**Appendix**). A third reviewer (PCD) was consulted in case of disagreement. For each included study, the following information was extracted: information on the exposure and outcome of interest; sample size; the genetic instrument; the MR design (one-sample and/or two-sample); population ancestry; and the MR analysis results. Following the Mendelian randomization dictionary ^20^, we defined one-sample MR studies as those we interpreted as measuring the instrumental variable-exposure and instrumental variable-outcome associations in the same sample ^21^, and two-sample Mendelian randomization studies as those which measured these associations in different samples ^22^. Inverse variance weighted models were regarded as the main analysis ^23,24^. Individual-level (usually associated with one-sample MR) and summary-level approaches (usually two-sample MR) can be conducted in the same population. We extracted both sets of results where studies explicitly reported both one- and two-sample results (e.g., Casanova et al 2021 ^25^).

### Quality assessment

We developed a scoring system incorporating factors relating to the validity of MR studies, that reflected the recently developed STROBE-MR guidelines ^19^. Important indices of quality are measurement of exposure and outcome (two items), selection of genetic variants (three items), results of MR analysis (five items for one-sample MR, seven items for two-sample MR). Each item was given a score of “***-***”, “***-+***”, or “***+***” corresponding to higher, moderate, or low risk of bias respectively. For example, regarding the type of measure for the exposure, we rated this “**-**” if unclear or self-reported, “***-+***” if a mix of self-reported and measured, and “***+***” if objectively measured. Additional details are presented in ***Supplementary material 3*** . Categories were given an overall score based on the most frequently occurring score, or the even score where two scores occurred at the same frequency as detailed in ***Supplementary material 4*** . For example, for “selection of genetic variants”, which was measured by three items, we chose the modal score (i.e., the score that appeared most frequently) to indicate quality of selection of genetic variants; for scores that occurred at the same frequency, we chose the higher rounded even score i.e., “**+**” was chosen between “**-+**” and “**+**”, “**-+**” was chosen between “**-+**” and”**-**”,“**+-**” was chosen between “***-***” and “***+***”. For MR studies that did not describe p-value threshold, we calculated this from the F statistic, % variance explained, or the variance from related MR studies that used similar/closely related sets of genetic variants as instruments. The lead reviewer (MG) and another reviewer (PJD, DAK, or YY) scored included studies independently, after which they compared their scores and any inconsistencies were resolved by discussion, or referral to a third reviewer (PCD).

### Statistical analysis

Most studies reported the odds ratio (OR) for a 1 standard deviation (SD) increase in adiposity indices, such as body mass index (BMI). Where this was not the case, we rescaled the units of the outcome and corresponding standard errors to 1 SD. Where the unit of exposure was not reported, we assumed it to be 1 SD, as this is how Mendelian randomization studies with continuous exposure typically report outcomes when using adiposity-related exposures. Exposures defined categorically were not included for meta-analyses due to their inherent limitations in capturing the full spectrum of exposure data and potentially introducing bias. Where the exposure-outcome was expressed as a regression coefficient for a continuous outcome, we used the Chinn equation to produce corresponding ORs to facilitate meta-analysis ^26^. Probit regression coefficients were converted into approximated logit coefficients by multiplying the probit coefficient by 1.6 ^27^. We found that many included papers used the same cohorts to derive the genetic instruments, so when more than one MR estimate was published on the same outcome and study population, the MR estimate based on the largest number of cases, or if cases were identical, the largest number of genetic variants was retained for meta-analysis. For outcomes reported as continuous or binary on the same scale (e.g., Patient Health Questionnaire-9) among the same cohort, number of cases, and number of genetic instruments we retained the binary clinical outcome for meta-analysis (similar to Larsson & Burgess, 2021 ^23^) (***Supplementary material 5)*.**

Random-effects models were used to produce summary effect sizes with 95% confidence intervals (CIs) from individual study data^28,29^. The percentage of variability in effect estimates that due to heterogeneity between studies was quantified using the I^2^ statistic. For this, values <25%, 25-75%, and >75% were considered to indicate low, moderate, and high heterogeneity respectively ^30^. Pre-specified subgroup analyses by sex were conducted if sex-specific estimates were provided. Where studies did not explicitly report ancestry, we examined the exposure and outcome genome-wide association studies (GWASs) to infer likely ancestry. Sensitivity analyses were conducted to investigate the equivalence of a meta-analysis of odds ratios and beta coefficients; thus, the main analysis was repeated including only studies reporting ORs with 95% CI and, separately, the main analysis with beta coefficients and 95% CI. All statistical analyses were conducted in Stata 14 SE (StataCorp, College Station, TX, USA) using the metan commands.

## Results

The search strategy resulted in 1,935 articles, of which 41 eligible Mendelian randomization studies with 184 estimates were included in this review (**Figure 1, *Supplementary material 1*** and ***Supplementary material 2***). Among them, nineteen studies reported 1 estimate ^31–49^, five studies reported 2 estimates ^50–54^, two studies reported 3 estimates ^55,56^, three studies reported 4 estimates^57–59^, and twelve studies reported 5+ estimates ^25,60–70^. Study characteristics and reported associations of the included studies are given in **Table 1**, and additional details are presented in ***Appendix***. Twenty-eight studies reported 98 estimates of the association between adiposity and depression ^25,32–35,37,40,42–45,47,48,50,53,55–59,62–64,66–70^, three studies reported 9 estimates for anxiety ^25,41,67^, three studies reported 8 estimates for anorexia nervosa ^61,67,68^, five studies reported 13 estimates for bipolar disorder ^31,53,55,65,67^, three studies reported 7 estimates for OCD ^61,67,68^, three studies reported 23 estimates for PTSD ^49,60,67^, and twelve studies reported 26 estimates for schizophrenia ^36,38,39,46,51,52,54,55,61,65,67,68^. One-sample MR design was conducted in six studies ^33,34,43,56,57,62^, two-sample MR design was conducted in thirty-two studies ^31,32,35–42,44–55,58,60,61,63–69^, both one-sample MR and two-sample MR were conducted in three studies ^25,59,70^. Thirty unique MR estimates out of 184 total estimates were included in meta-analyses (details as shown in “Yes” under “For meta-analysis” collum in Appendix).

**Table 1.**
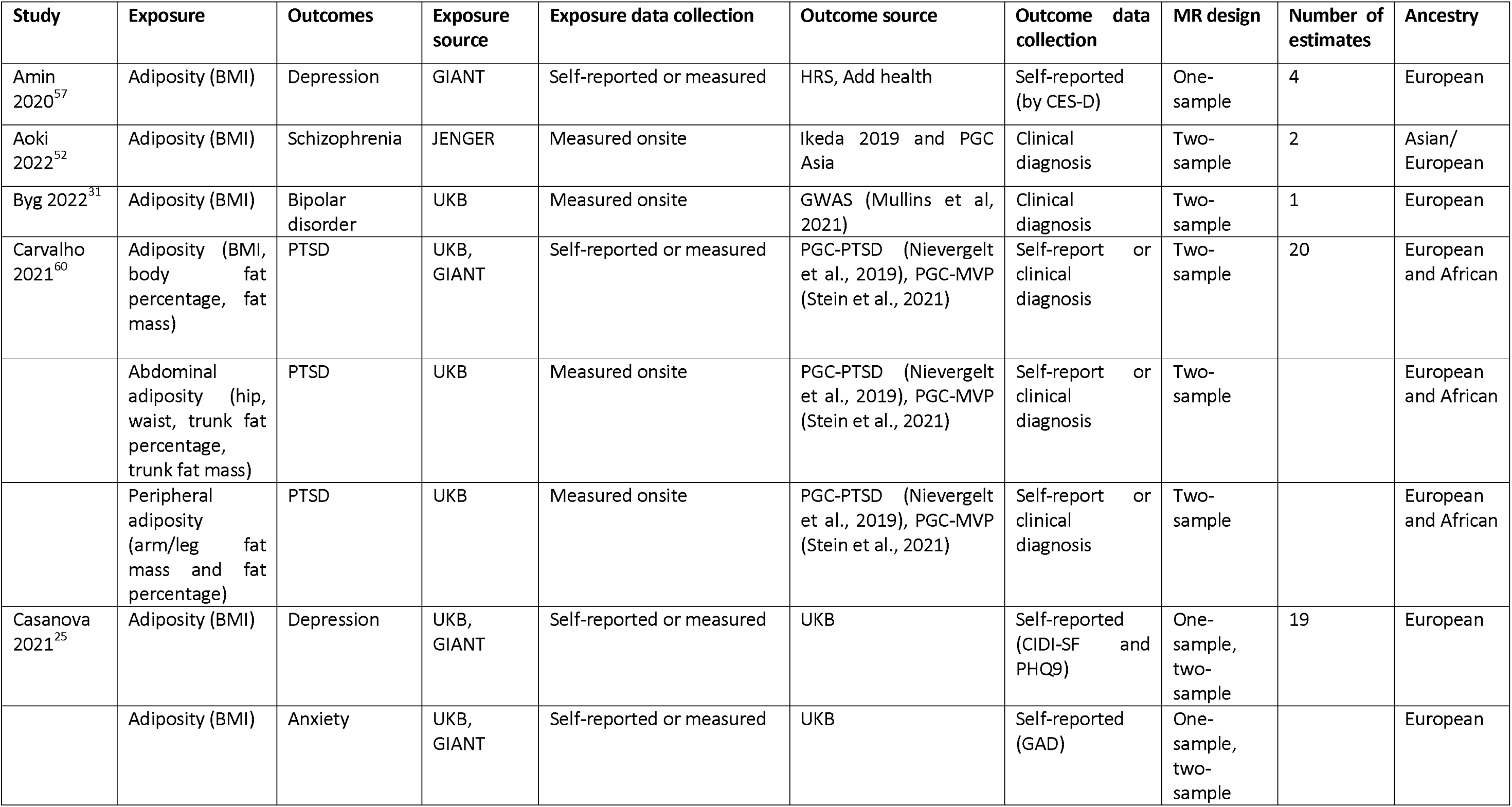
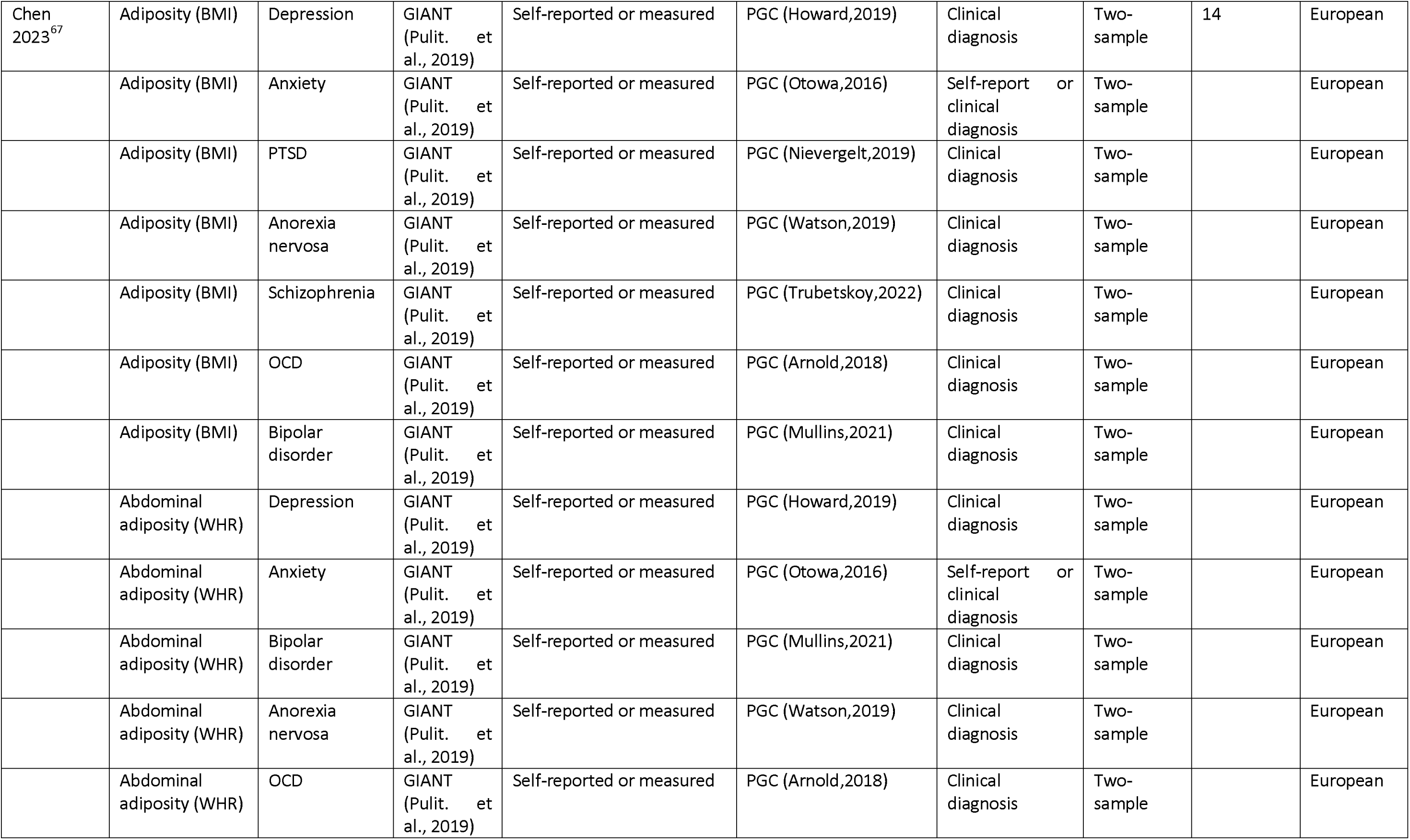
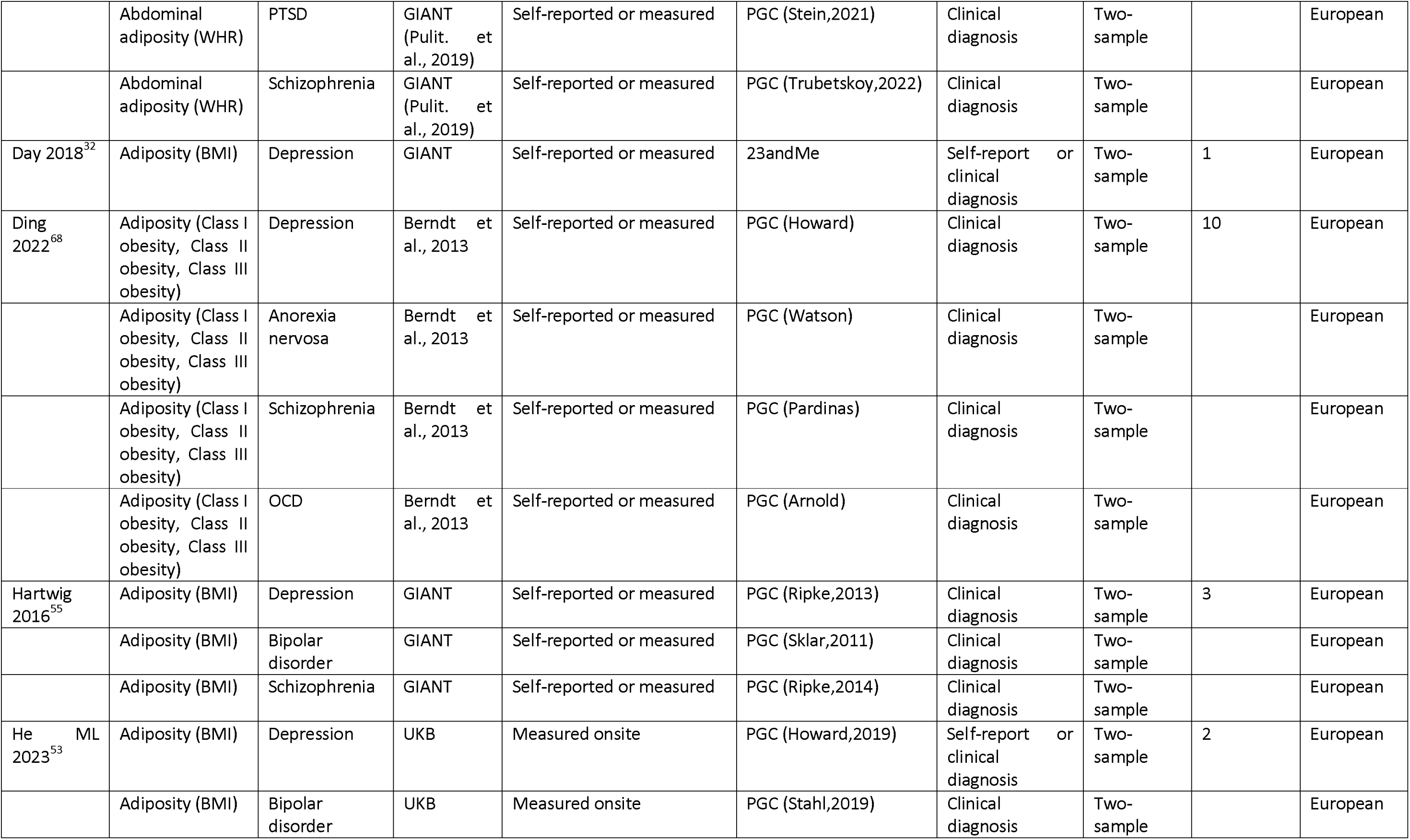
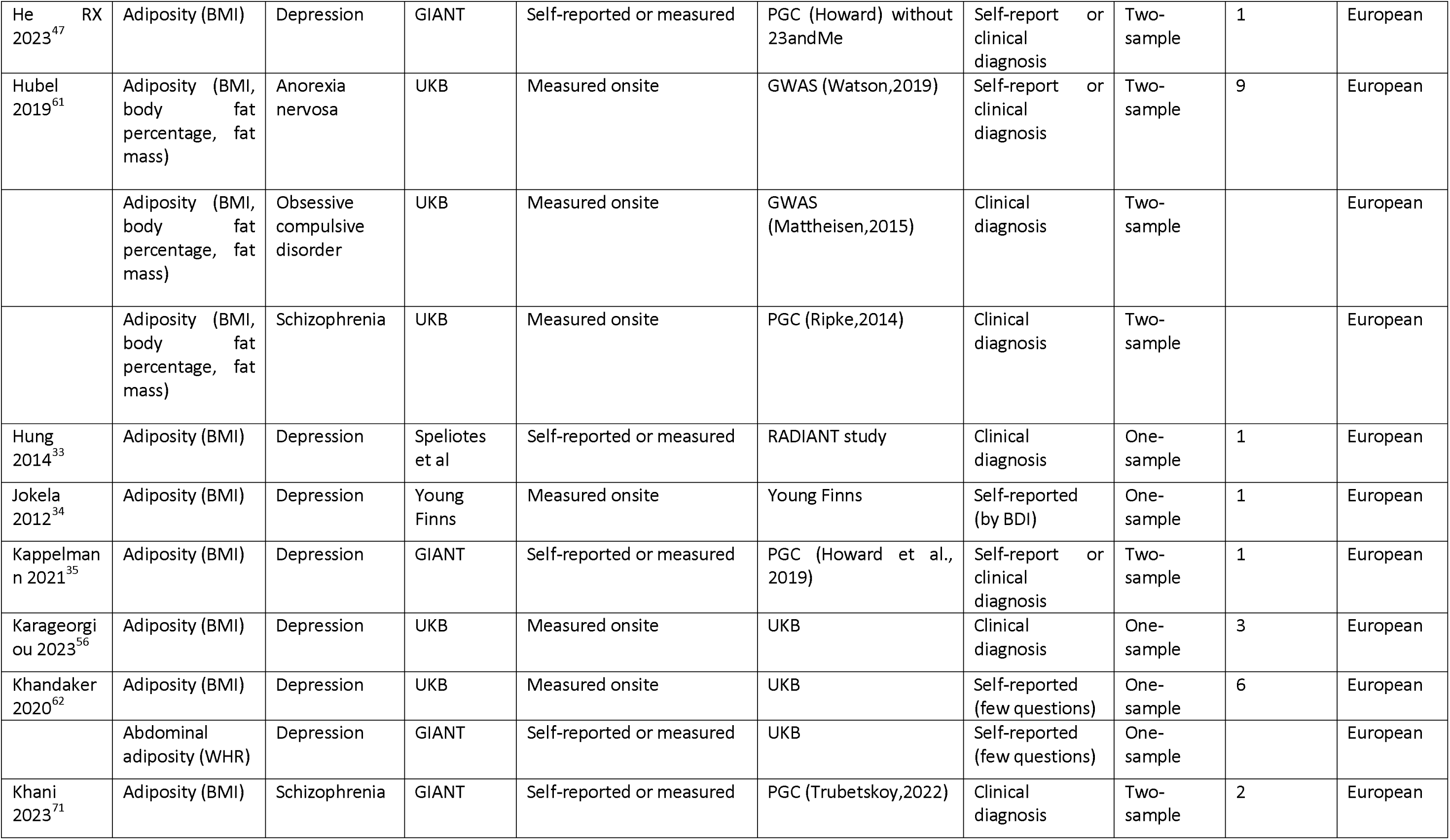
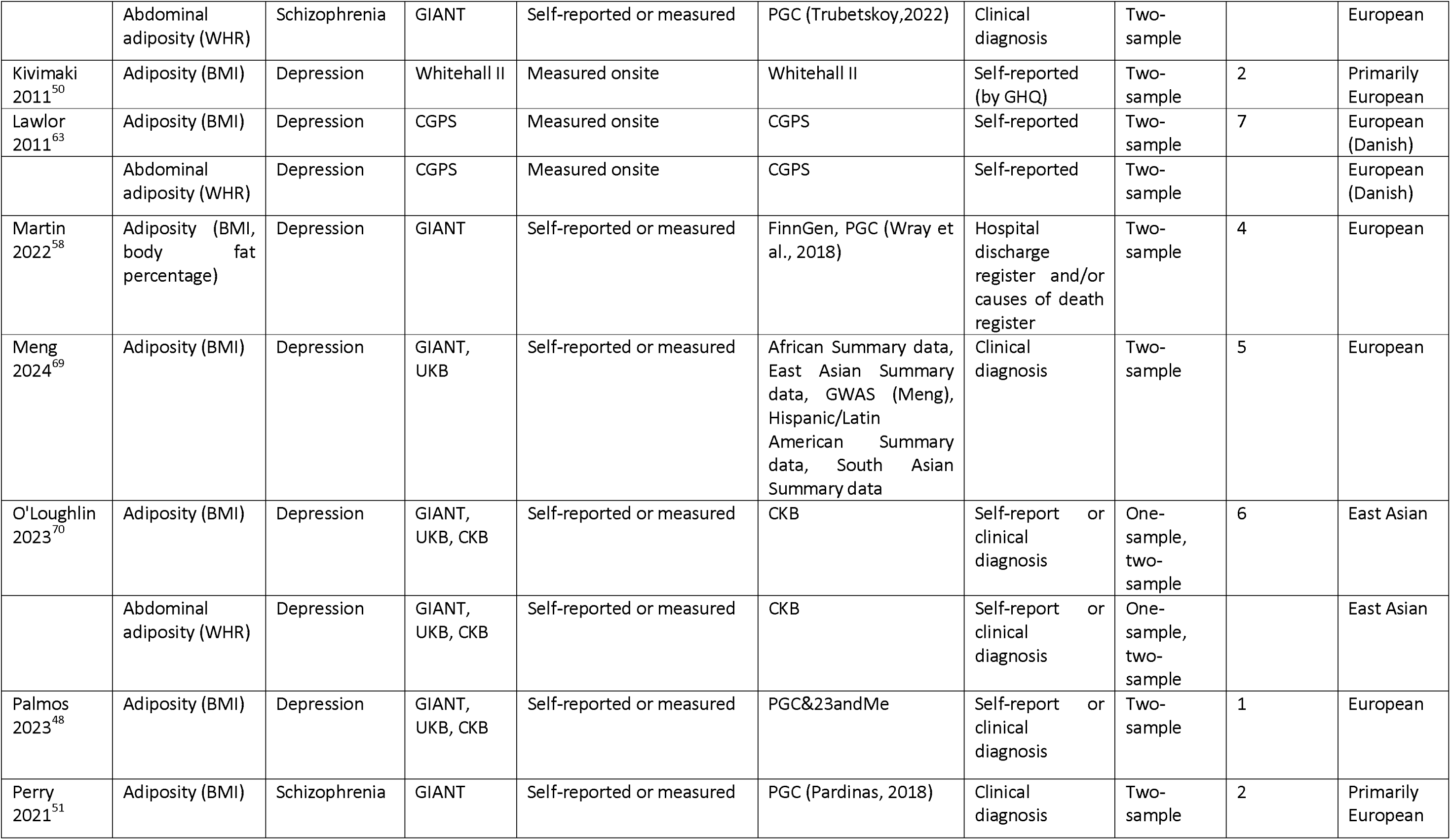
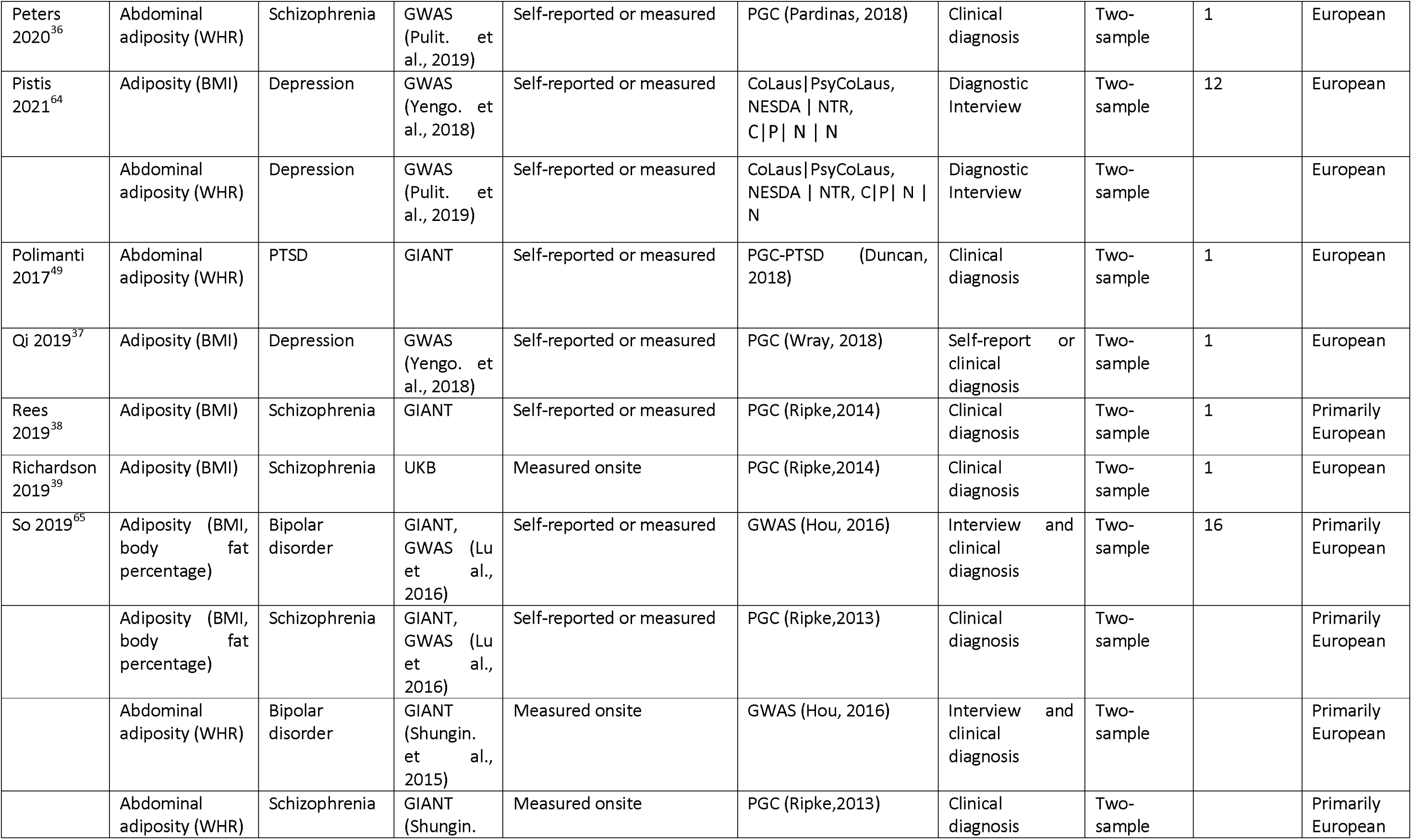
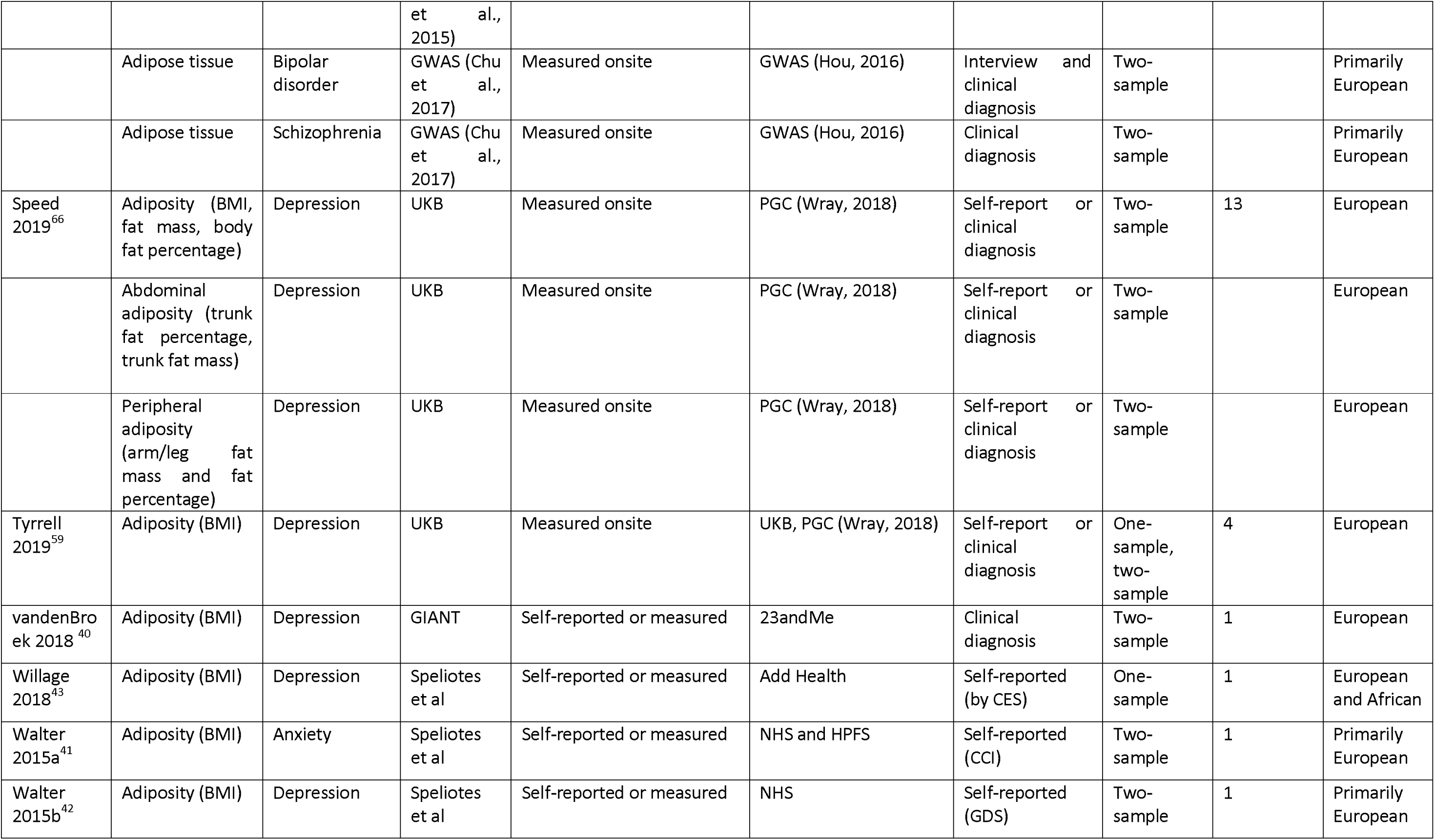
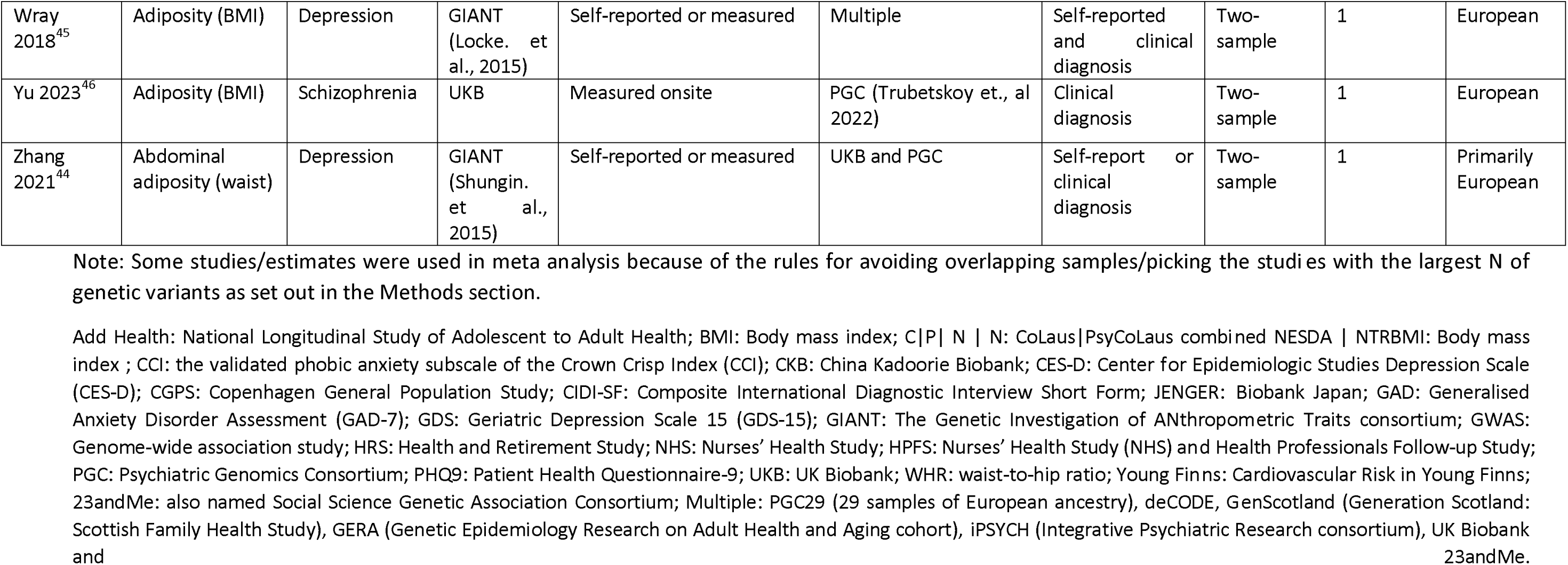
The characteristics of included Mendelian randomization studies of adiposity measures on mental illnesses.

### Quality assessment of included studies

The quality assessment of studies included in this review is presented in **Table 2** and ***Supplementary material 4*** . For the assessment of exposure and outcome, 86% of studies used adiposity data from the UK Biobank and mental illness data from Psychiatric Genomics Consortium. For the selection of genetic variants, most studies selected a genetic variant p-value threshold below 5E-08 (n=32 studies)^25,31,32,35,37–40,43–49,51–61,64–70^, but an F-statistic ≥10 (n=27 studies) ^25,32–36,38,44,47–49,51,53,55–60,62–65,67,68,70^, and had a high percentage (≥3%) of explained variance (n=11 studies) ^32,35,38,51,55,58,60,62,64,66^, providing good evidence that the chosen instrument was robust.

**Table 2.** Methodological quality assessment of the included Mendelian randomization studies *.

| Author | Measurement of exposure and outcome | Selection of genetic variants | Analysis (One-sample MR) | Analysis (Two-sample MR) |
| --- | --- | --- | --- | --- |
| Amin 2020<sup>57</sup> | - + | + | - |  |
| Aoki 2022<sup>52</sup> | + | - |  | + |
| Byg 2022<sup>31</sup> | + | - + |  | + |
| Carvalho 2021<sup>60</sup> | - + | + |  | + |
| Casanova 2021<sup>25</sup> | - + | + | - | + |
| Chen 2023<sup>67</sup> | + | + |  | + |
| Day 2018<sup>32</sup> | - + | + |  | + |
| Ding 2022<sup>68</sup> | + | + |  | + |
| Hartwig 2016<sup>55</sup> | + | + |  | + |
| He ML 2023<sup>53</sup> | + | + |  | + |
| He RX 2023<sup>47</sup> | - + | + |  | + |
| Hubel 2019<sup>61</sup> | + | - |  | + |
| Hung 2014<sup>33</sup> | + | - + | - |  |
| Jokela 2012<sup>34</sup> | + | - + | - |  |
| Kappelmann 2021<sup>35</sup> | - + | + |  | + |
| Karageorgiou 2023<sup>56</sup> | + | + | + |  |
| Khandaker 2020<sup>62</sup> | - + | + | - |  |
| Khani 2023<sup>54</sup> | + | - |  | + |
| Kivimaki 2011<sup>50</sup> | + | - |  | - |
| Lawlor 2011<sup>63</sup> | - + | - |  | - |
| Martin 2022<sup>58</sup> | + | + |  | + |
| Meng 2024<sup>69</sup> | - + | - |  | + |
| O'Loughlin 2023<sup>70</sup> | - + | + | + | + |
| Palmos 2023<sup>48</sup> | - + | + |  | + |
| Perry 2021<sup>51</sup> | + | + |  | + |
| Peters 2020<sup>36</sup> | + | - + |  | + |
| Pistis 2021<sup>64</sup> | + | + |  | + |
| Polimanti 2017<sup>49</sup> | + | + |  | + |
| Qi 2019<sup>37</sup> | - + | - |  | + |
| Rees 2019<sup>38</sup> | + | + |  | + |
| Richardson 2019<sup>39</sup> | + | - |  | + |
| So 2019<sup>65</sup> | + | + |  | + |
| Speed 2019<sup>66</sup> | + | + |  | + |
| Tyrrell 2019<sup>59</sup> | + | + | - | + |
| vandenBroek 2018<sup>40</sup> | + | - + |  | + |
| Walter 2015a<sup>41</sup> | - + | - |  | - |
| Walter 2015b<sup>42</sup> | - + | - |  | - |
| Willage 2018<sup>43</sup> | - + | - | - |  |
| Wray 2018<sup>45</sup> | - + | - |  | + |
| Yu 2023<sup>46</sup> | + | + |  | + |
| Zhang 2021<sup>44</sup> | - + | - |  | + |

Note: Green colour indicates low risk of bias, yellow colour indicates moderate risk of bias, pink colour indicates high risk of bias, grey colour indicates not applicable.

Among the 35 studies using two-sample MR design, 31 studies scored “+” in the analysis category^25,31,32,35–40,44–49,51–55,58–61,64–70^, and only four studies scored “-” in the analysis category ^41,42,50,63^, meaning that most studies tested the majority of: bidirectional effects, non-linearity assessed for continuous exposures, heterogeneity, reported data on pleiotropy-robust sensitivity analysis, similar results amongst all MR analyses, and for two-sample MR SNP harmonization across samples, and estimates SNP-exposure and SNP-outcome from comparable populations. Among the nine included studies using a one-sample MR design, two studies scored “+” in the analysis category ^56,70^, with the remaining seven studies scoring “-” ^25,33,34,43,57,59,62^.

### Meta-analyses results for general adiposity and mental illnesses

Thirty-eight studies with 123 estimates provided data for the association between general adiposity and mental illnesses ^25,31–35,37–43,45–48,50–70^, and sixteen studies with 23 estimates were included in meta-analysis ^25,33,34,41,42,46,50,52,57,58,60,63,64,67,69,70^, as shown in **Figure 2A** and **Figure 2B** . Most studies presented data for the association with depression. There was generally consistent, but imprecise, evidence that genetically predicted higher adiposity was associated with an increased risk of depression, with a combined OR of 1.07 (95% CI 1.00-1.13, n=16 estimates) per SD of general adiposity, but with high heterogeneity (*I*^2^ = 83.4%) between studies. However, there was no evidence of a casual association between genetically predicted adiposity and anxiety (OR 1.12, 95% CI 0.97-1.29, n=3 estimates) ^25,41,67^, PTSD (OR 1.15, 95% CI 0.87-1.54, n=2 estimates) ^60,67^, schizophrenia (OR 0.95, 95% CI 0.73-1.23, n=2 estimates) ^46,52^. Most of these analyses showed high heterogeneity.

### Meta-analyses results of abdominal adiposity with mental illnesses

Twelve studies with 12 estimates reported the causal relationship between abdominal adiposity and mental illness ^36,44,49,54,60,62–67,70^, and 5 studies with 7 estimates were included in meta-analyses after excluding overlapped study populations ^60,63,64,67,70^, as shown in **Figure 3**. There was no evidence of a causal association between abdominal adiposity and depression (OR 0.95, 95% CI 0.84-1.08, n=5 estimates) ^63,64,67,70^ or abdominal adiposity and PTSD (OR 1.12, 95% CI 0.93-1.34, n=2 estimates) ^60,67^.

### Description of the associations between adiposity indices and other mental illnesses

One study with 6 estimates on the associations of adipose tissue and bipolar disorder and schizophrenia was not included for meta-analyses because estimates were derived from a single cohort (**Appendix**)^65^. For general adiposity, five studies examined the relationship with bipolar disorder, but their study cohorts overlapped, with the largest study reporting a risk of bipolar disorder of 0.96 (95% CI 0.89-1.03)^67^. Similar overlaps were observed for the association between general adiposity and anorexia nervosa, with the largest study reporting a risk of 1.43 (95% CI 1.30-1.58)^67^, and for associations between general adiposity and OCD with the largest study reporting a risk of 0.68 (95% CI 0.54-0.85)^67^. For peripheral adiposity, two studies with 17 estimates reported the casual relationship between peripheral adiposity and mental illnesses ^60,66^. Associations for major depression were not included in the meta-analysis due to overlapped cohorts. However, most studies demonstrated a significant causal association. Associations for PTSD were not included in the meta-analysis because this study only reported significant results from European ancestry, results from African ancestry were eligible but not included in the meta-analysis due to overlapped cohorts. For abdominal adiposity, one study reported the associations of abdominal adiposity with anxiety (1.23 [95% CI 0.98-1.56]), anorexia nervosa (1.29 [95% CI 1.15-1.45]), and OCD (0.59 [95% CI 0.44-0.78])^67^.

### Subgroup analyses

A pre-specified subgroup analysis was performed to estimate the associations of genetically predicted adiposity measures with mental illnesses stratified by sex. In **Appendix**, seven studies with 76 estimates provided the associations of genetically predicted BMI with depression and anxiety by sex ^25,43,57,59,70^. In **Figure 4A**, there is no evidence from a meta-analysis including 18 MR estimates that the causal relationship between BMI and depression differed by sex (P=0.313). Similar results were also found for anxiety (P=0.287) as shown in **Figure 4B**^25,41^.

### Sensitivity analyses for clinical diagnosis of depression

We repeated the main analyses estimating the association of genetically predicted adiposity with depression including only studies that reported results with binary outcomes. In **Appendix**, nine studies with 15 estimates provided results with binary outcomes (OR 1.06, 95% CI 1.00-1.13) ^33,50,57,58,63,64,67,69,70^. The findings were similar to the main analysis which included binary and continuous outcomes (**Figure 5**).

## Discussion

This systematic review and meta-analysis of Mendelian randomization studies regarding adiposity and mental illnesses found a causal relationship between general adiposity and depression, although effect sizes were modest. No sex difference was observed in this causal relationship. Evidence was uncertain for the association between different measures of adiposity and PTSD, schizophrenia, bipolar disorder, anorexia nervosa, and OCD. The findings from this review and meta-analysis are timely and address an important evidence gap. Presently, 39% and 12.5% of adults worldwide live with obesity and mental illness respectively ^72,73^, our findings suggests that higher general adiposity can cause depression.

A systematic review of 8 MR studies on obesity and depression with a last literature search in October 2021 found consistent evidence, with a higher depression risk associated with per SD in BMI (OR:1.33, 95% CI 1.19-1.48) ^74^. Compared to that review, we selected studies with the largest GWAS data available for more types of mental illnesses and avoided selecting the same or overlapping cohorts where larger studies were available. This accounts for a large proportion of MR associations that were not included for meta-analysis. For example, we selected studies that utilized the PGC (Howard) dataset for depression over those using the PGC (Wray) dataset (***Supplementary material 5)***. Our results are in line with a meta-analysis of randomized trials of behavioral weight loss programs found modest improvements in depression at 12 months from baseline, but with insufficient data for symptoms of other mental illnesses ^75^.

Apart from depression, the causal effect of adiposity on other mental illnesses has been less studied using MR study designs, and findings are less certain but do not exclude meaningful causal associations. Meta-analyses from conventional epidemiological research designs reported that obesity is associated with an increased likelihood of anxiety disorder (OR 1.30, 95% CI 1.20-1.41) ^76^, and an increased prevalence of bipolar disorder (OR 1.77, 95% CI 1.40-2.23) ^77^. In our meta-analyses, we did not find evidence of a causal role of adiposity in anxiety, PTSD, bipolar disorder, schizophrenia, anorexia nervosa, or OCD, but data were imprecise, so more data would aid in further elucidating this relationship. Similarly, the small number of MR studies that assessed evidence of a causal relationship specifically between abdominal adiposity and mental illnesses, including depression, produced imprecise evidence compatible with either no risk or moderate-sized increased risks. Conventional study designs have indicated that fat distribution (abdominal adiposity vs general adiposity) is associated with depressive symptoms independent of general adiposity ^78^. Given the small number of MR studies that reported abdominal adiposity and mental illness, further MR research with different and larger samples is needed to explore the causality of abdominal adiposity for mental illnesses.

Assessing the quality of analyses in MR is challenging due to the complexity and interrelationships between different elements of study design and the type of analysis used. A limitation of our review is that we attempted to characterize some of the main sources of possible bias, but arguably did not characterize all potential sources. For example, it was difficult, given reporting standards in some of the included literature, to fully characterize any sample overlap and the extent to which this may have biased two-sample analysis. A similar issue arose with characterizing the genetic ancestries included in each study. To guide our quality assessment, we developed a checklist that reflected STROBE MR guidance. Using this quality assessment tool, we observed differences in methodological quality between the included studies. We focused on potential violations of the instrumental variable assumptions that underlie MR analyses, particularly the quality of analysis reported in each study. For example, a critical issue that may introduce bias is violations of the exclusion restriction due to pleiotropy. Newer studies tended to perform better when assessed against criteria relating to the quality of analysis, whereas older studies were undertaken before methods for pleiotropy sensitivity analysis were fully developed.

This review has several other limitations. First, statistical heterogeneity in most meta-analyses was moderate to high. This suggests that more evidence may be needed to investigate or strengthen the causal association of more indices of adiposity with mental illness. Second, selection bias should be considered. For example, participants from cohorts such as the UK Biobank are relatively healthier than the general population and studies relying on evidence from volunteer cohorts of healthy and relatively affluent populations may be affected by collider bias associated with self-selection ^79^. Third, although this analysis provides evidence for the causal relationship between general adiposity and depression, we were unable to assess the mechanisms that may underlie this causal relationship. Fourth, most studies pertained to populations of predominantly European ancestry. Fifth, as noted, our quality assessment did not capture all aspects of potential bias in MR studies. For example, we did not identify studies that relied wholly on within-family designs, which may provide more robust estimates of causal associations in the presence of dynastic biases and cryptic population stratification ^80^. We did not formally assess studies for whether they relied on samples of unrelated individuals, and this appears to be an area where reporting could be improved in future MR research.

## Conclusions

Adiposity appears to be causally associated with depression. Evidence for associations with other mental health phenotypes remains unclear. Future studies are needed to elucidate the mechanistic causal links between adiposity and depression.

## Data Availability

All data produced in the present work are contained in the manuscript

## Data sharing

All the data supporting the conclusions of this article are included within the article and its supplementary files.

## Author contributions

**MG:** Conceptualization, Methodology, Software, Formal analysis, Investigation, Writing - Original Draft, Writing - Review & Editing. **PJD**: Methodology, Investigation, Writing - Review & Editing. **DAK**: Methodology, Investigation, Writing - Review & Editing. **YY**: Investigation, Writing - Review & Editing. **PA**: Conceptualization, Methodology, Writing - Review & Editing. **RS**: Statistical Methodology, Writing - Review & Editing. **PCD**: Methodology, Investigation, Writing - Review & Editing, Supervision. **MG** and **PCD** are guarantors of the study.

## Acknowledgements

We thank Nia Roberts for helping with the search strategy and the participants and investigators of all the cohorts and consortiums included in our study.

---

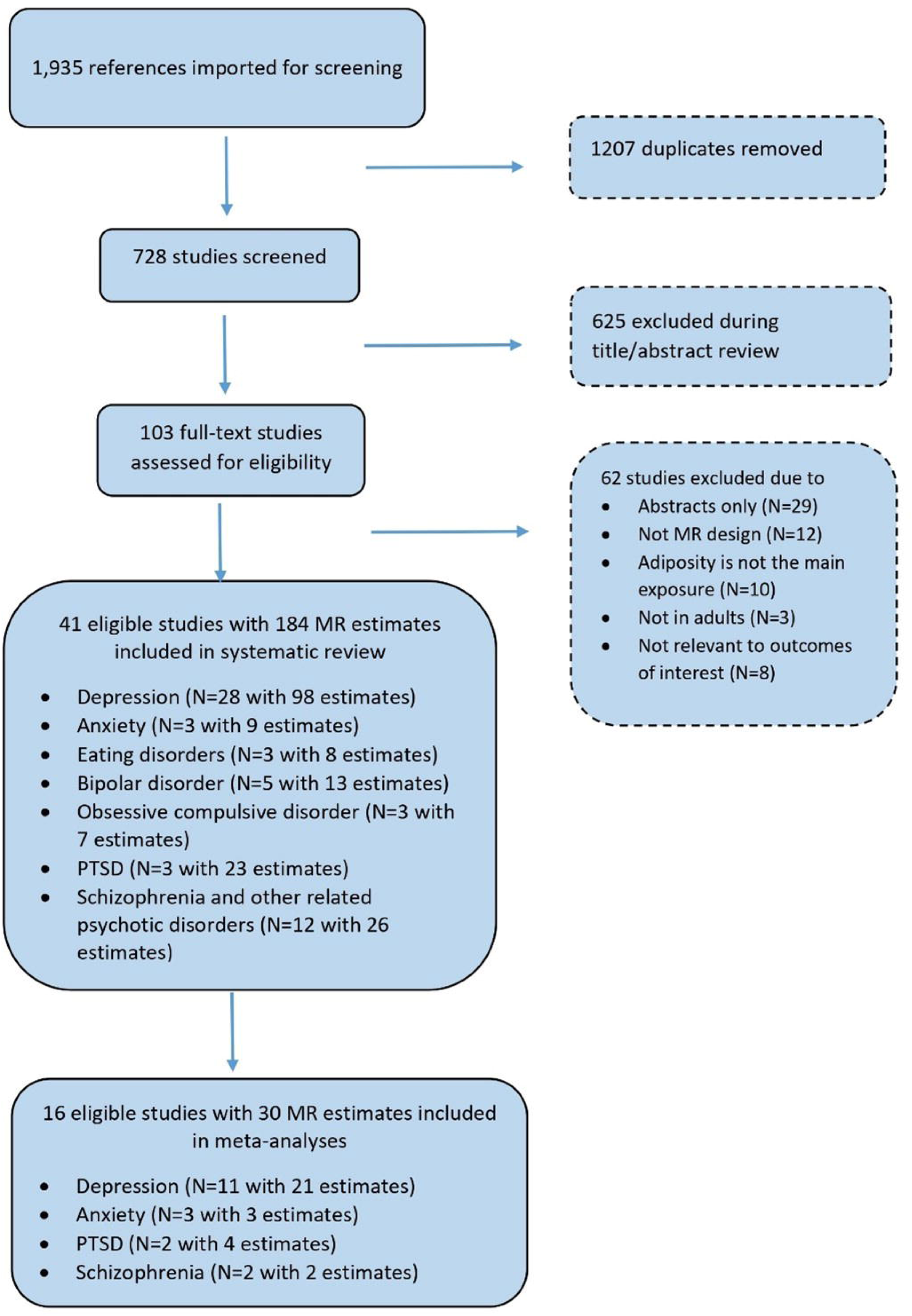

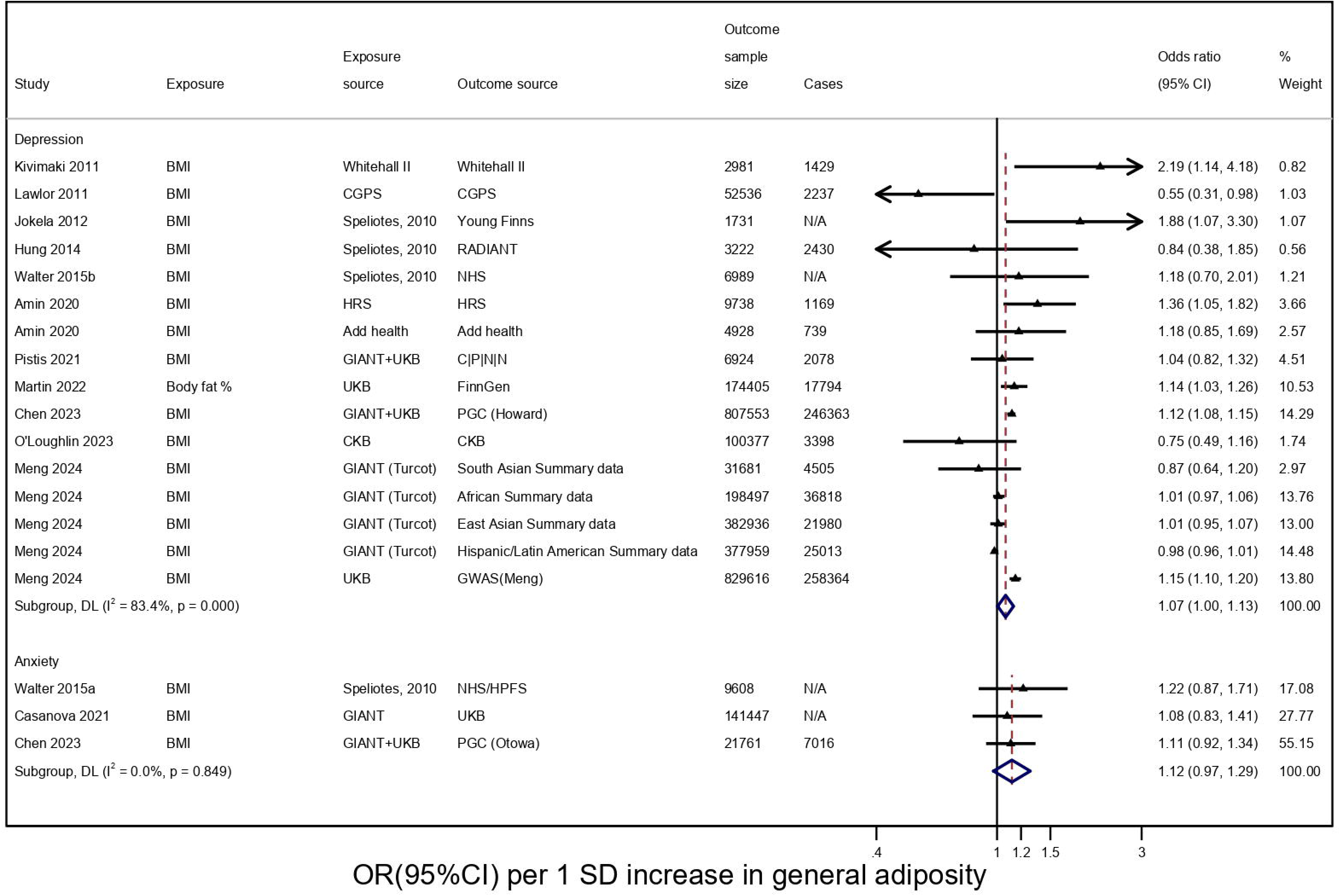

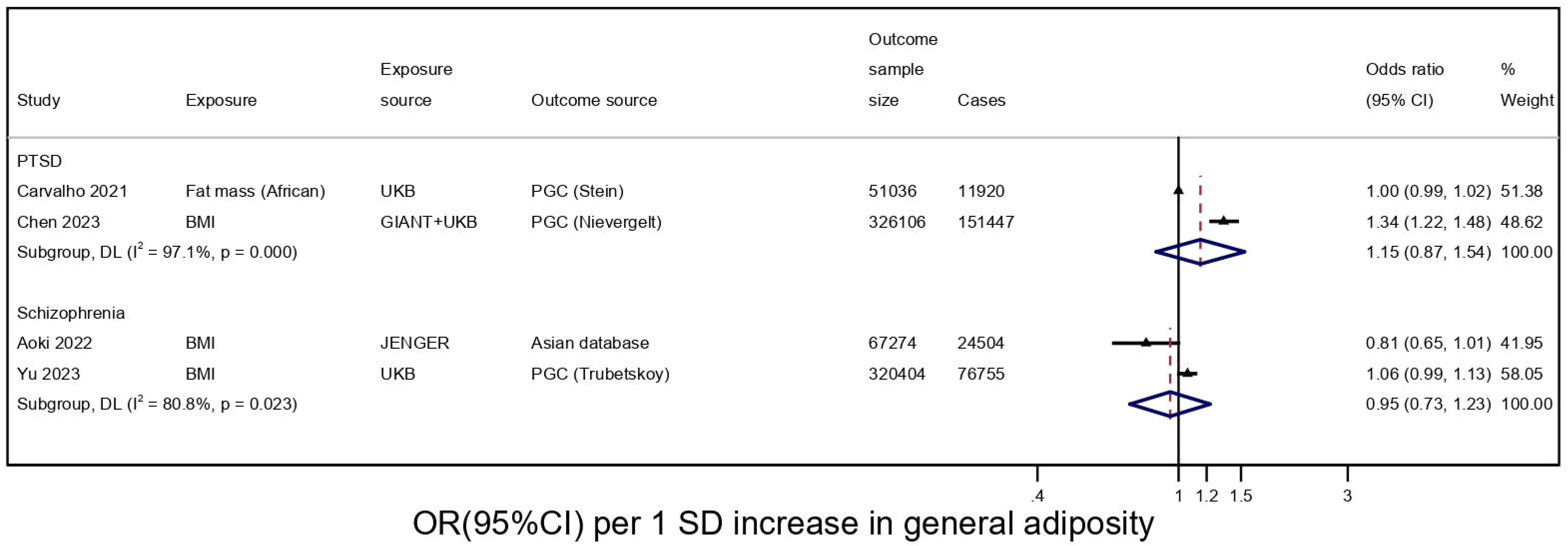

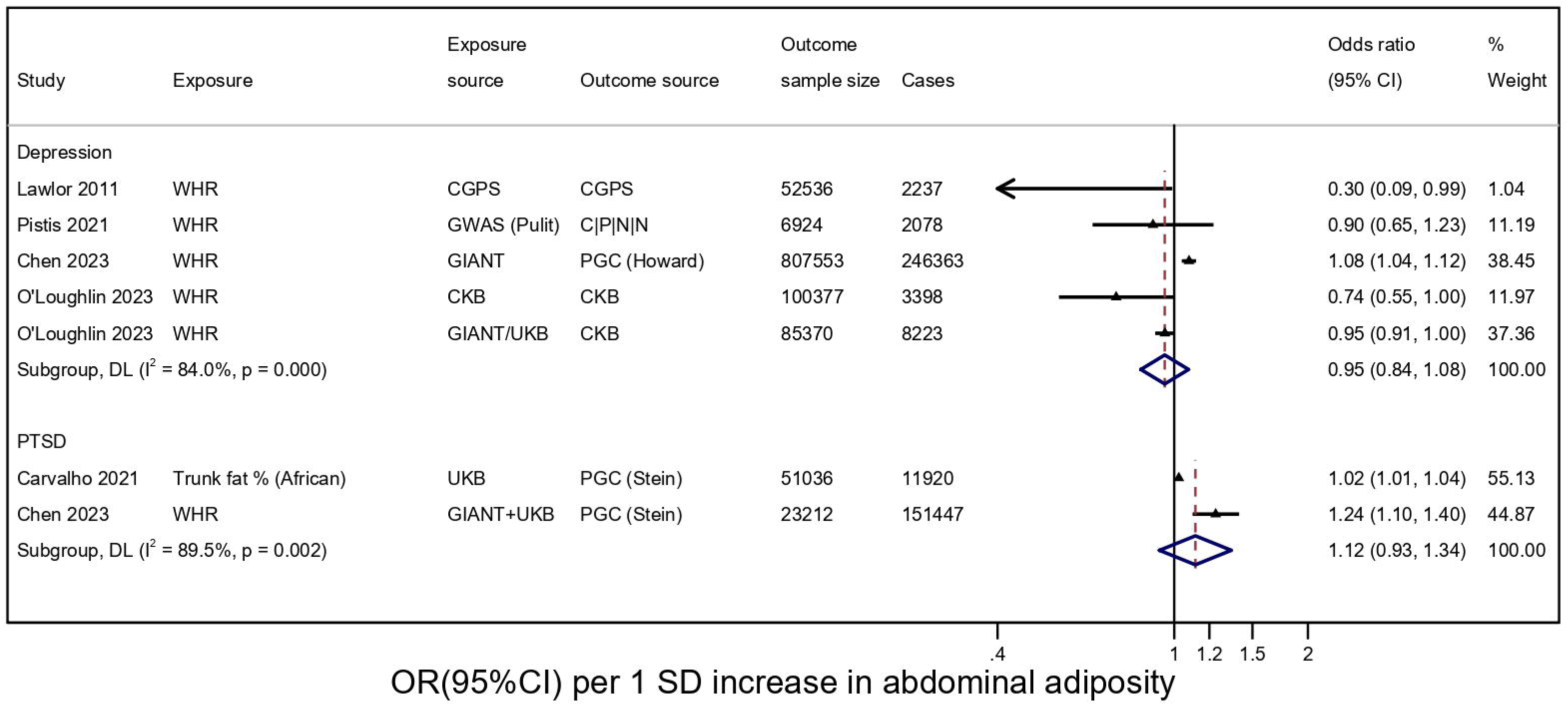

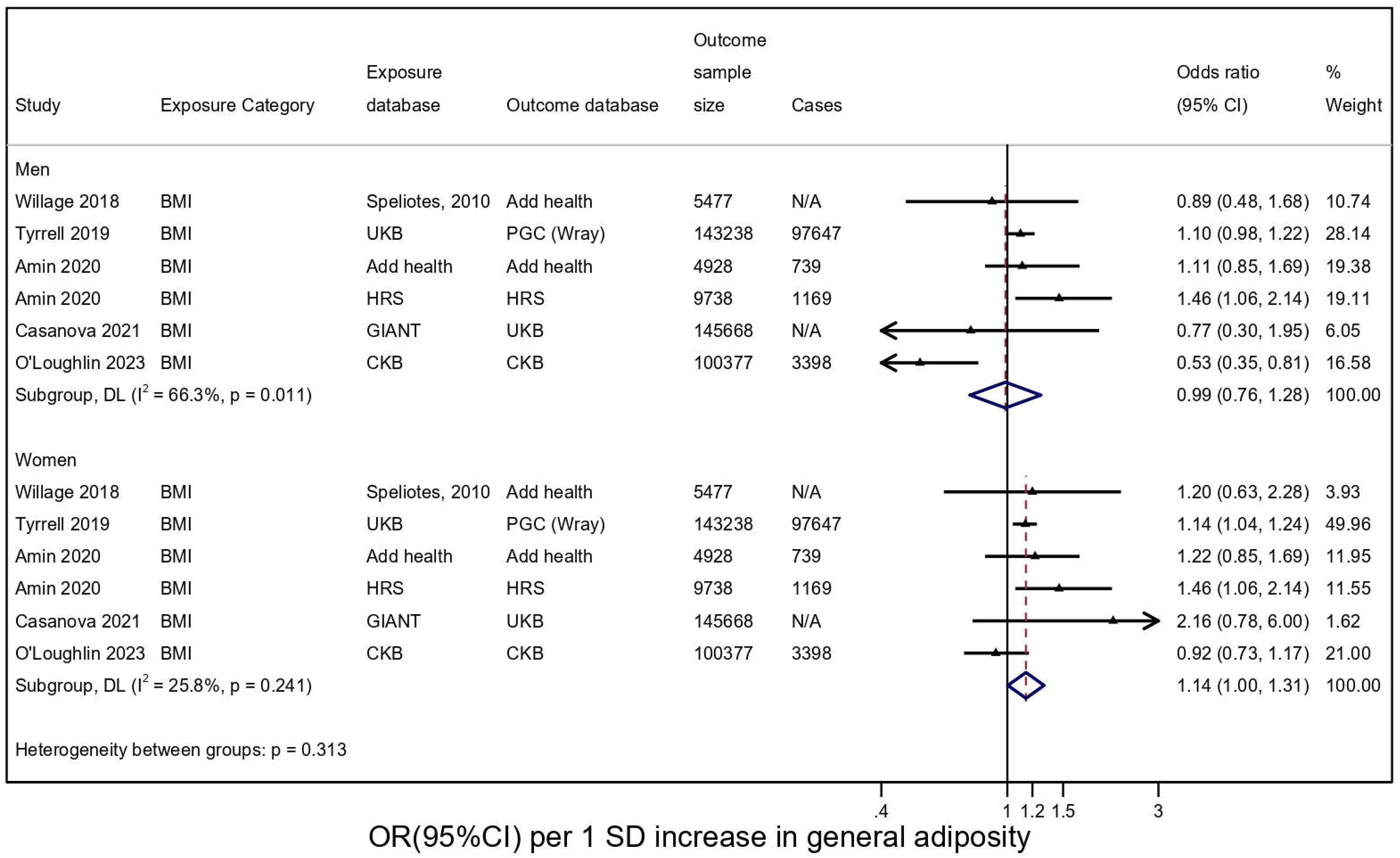

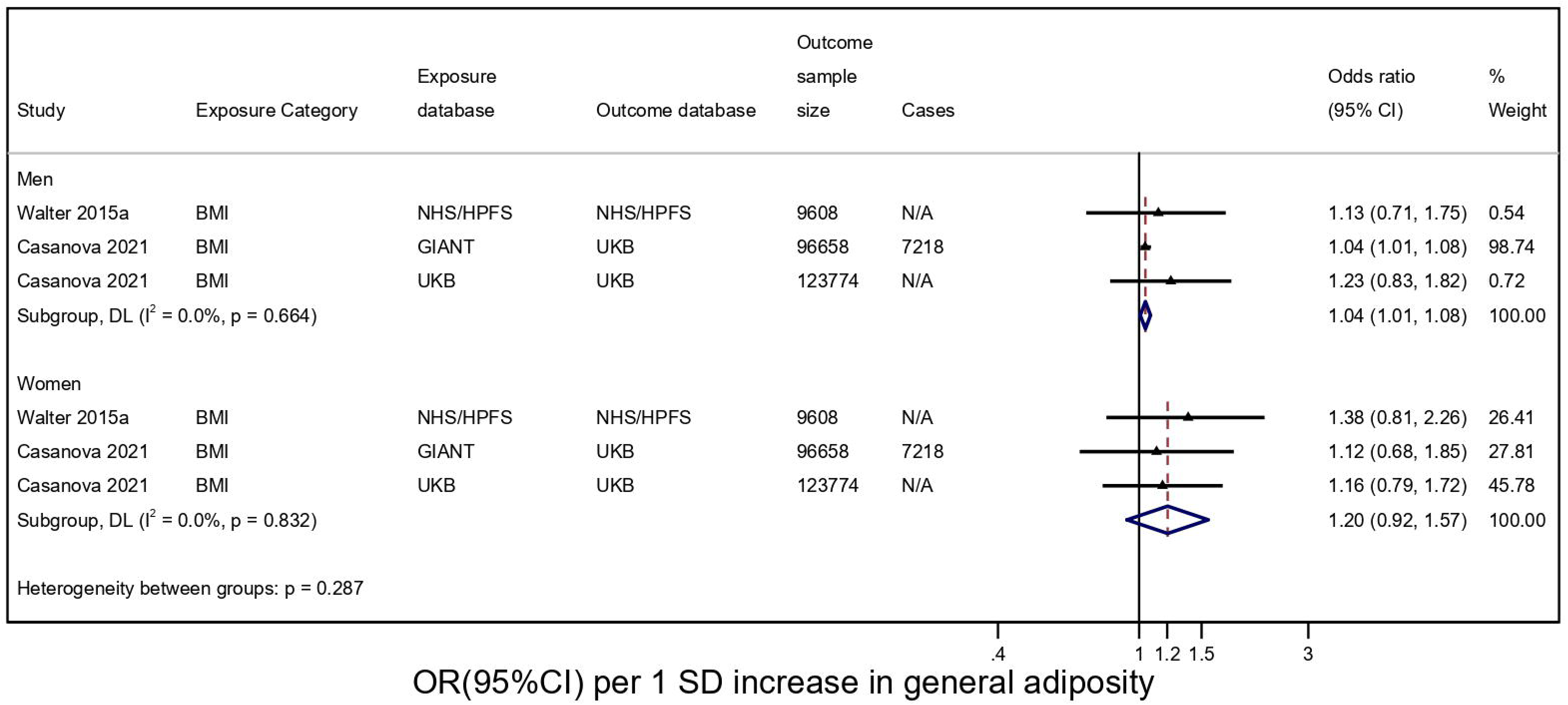

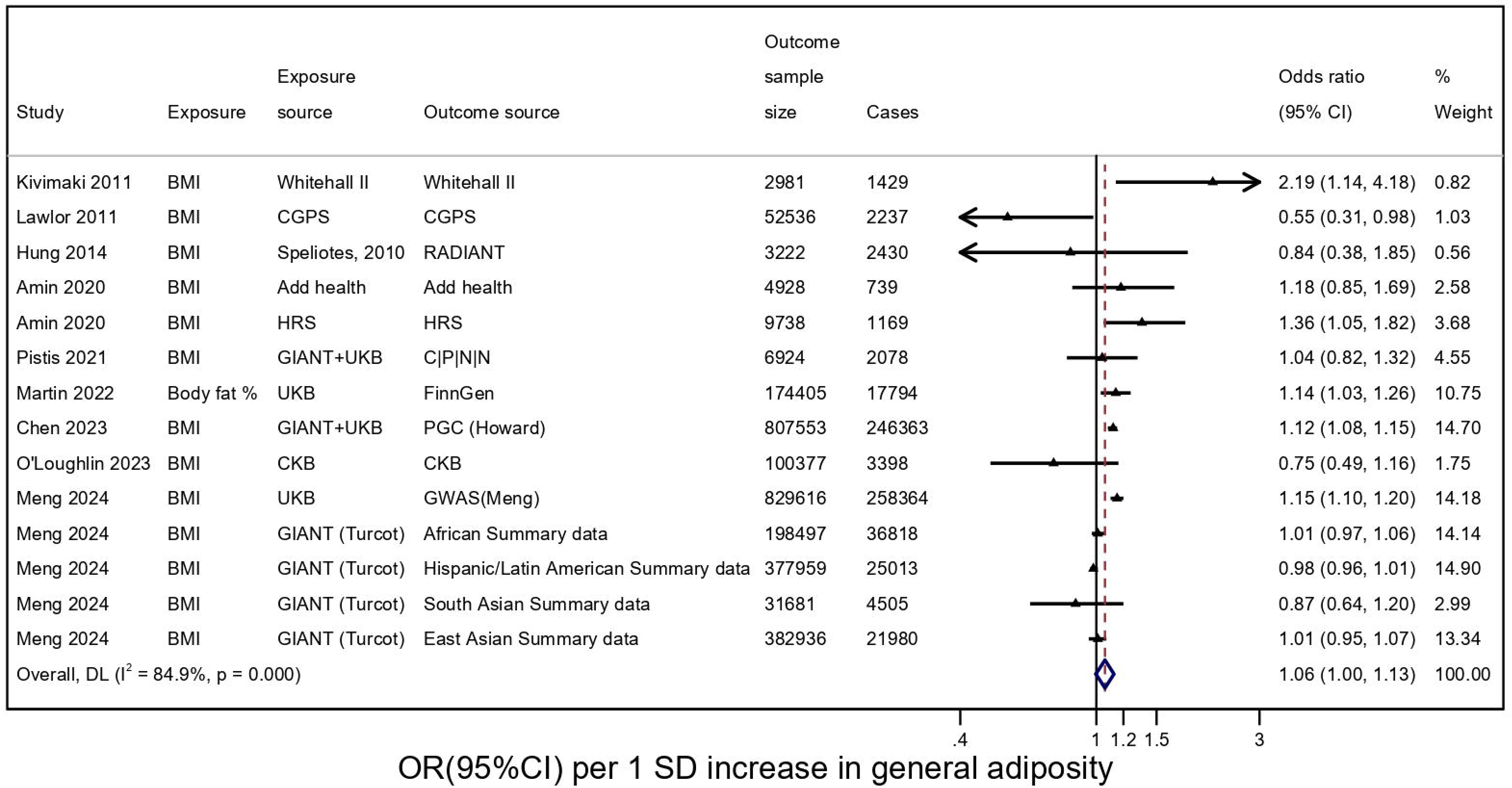

